# The 3p21.31 genetic locus promotes progression to type 1 diabetes through the CCR2/CCL2 pathway

**DOI:** 10.1101/2021.08.01.21261449

**Authors:** Paul Tran, Sharad Purohit, Eileen Kim, Khaled bin Satter, Diane Hopkins, Kathleen Waugh, Fran Dong, Suna Onengut-Gumuscu, Stephen S. Rich, Marian Rewers, Jin-Xiong She

**Affiliations:** Center for Biotechnology and Genomic Medicine, Medical College of Georgia, 1120 15^th^ Street, Augusta, GA, 30912, USA; Department of Obstetrics and Gynecology, Medical College of Georgia, Augusta University 1120 15^th^ Street, Augusta, GA, 30912, USA; Department of Undergraduate Health Professionals, College of Allied Health Sciences, Augusta University 1120 15^th^ Street, Augusta, GA, 30912, USA; Center for Public Health Genomics, University of Virginia School of Medicine, Charlottesville, VA, USA; Barbara Davis Center for Diabetes, University of Colorado Denver, Mail Stop A-140, 1775 Aurora Court, Aurora, CO 80045, USA

**Keywords:** CCR2, CCL2, MCP1, T1D, DAISY, autoimmune, fine mapping

## Abstract

**Objective:** Multiple cross-sectional and longitudinal studies have shown that serum levels of the chemokine ligand 2 (CCL-2) are associated with type 1 diabetes (T1D), although the direction of effect differs. We assessed CCL-2 serum levels in a longitudinal cohort to clarify this association, combined with genetic data to elucidate the regulatory role of CCL-2 in T1D pathogenesis.

**Research design and methods:** The Diabetes Autoimmunity Study in the Young (DAISY) followed 310 subjects with high risk of developing T1D. Of these, 42 became persistently seropositive for islet autoantibodies but did not develop T1D (non-progressors); 48 did develop T1D (progressors). CCL-2 serum levels among the three study groups were compared using linear mixed models adjusting for age, sex, HLA genotype, and family history of T1D. Summary statistics were obtained from the CCL-2 protein quantitative trait loci (pQTL) and *CCR2* expression QTL (eQTL) studies. The T1D fine mapping association data were provided by the Type 1 Diabetes Genetics Consortium (T1DGC).

**Results:** Serum CCL-2 levels were significantly lower in both progressors (p=0.004) and non-progressors (p=0.005), compared to controls. Two SNPs (rs1799988 and rs746492) in the 3p21.31 genetic locus, which includes the CCL-2 receptor, *CCR2*, were associated with increased *CCR2* expression (p = 8.2e-5 and 5.2e-5, respectively), decreased CCL-2 serum level (p = 2.41e-9 and 6.21e-9, respectively), and increased risk of T1D (p = 7.9e-5 and 7.9e-5, respectively).

**Conclusions:** The 3p21.31 genetic region is associated with developing T1D through regulatory control of the CCR2/CCL2 immune pathway.

## INTRODUCTION

Type 1 diabetes (T1D) is an autoimmune disorder where some genetically predisposed individuals are exposed to an environmental trigger which leads to immune recognition and destruction of pancreatic islet cells^1^. T1D is associated with microvascular and macrovascular complications and a shortened lifespan^2,3^. A primary goal in T1D is primary prevention of the disease. Recent success in anti-CD3 inhibition delaying progression from islet autoantibody positivity to T1D has brought promise to immunomodulation for T1D prevention^4^. The use of genome wide association studies (GWAS) may help identify other immune targets^5,6^.

GWAS have associated the 3p21.31 locus with T1D and pediatric autoimmune diseases (including T1D)^6,7^; however, the causal variants and mechanism of association have not been established for this locus. This region of the human genome is enriched for chemokine receptors that play an important role in autoimmunity^8^, including the C-C motif chemokine receptors (*CCR1, CCR2, CCR3, CCR5, CCR9, CCRL2, CXCR6*) and X-C motif chemokine receptor 1 (*XCR1*). Associated pathways include chromatin remodeling genes (*SETD2, ELP6*, and *SMARCC1*), putative transcriptional regulators (*CCDC12, CCDC51, CCDC36*, and *ZNF589*), and cell replication associated genes (*KIF9* and *CDC25A*)^9^, as well as the *CCRL2*-*LINC02009*^6^ and *DAG1*^7^ loci.

CCR2 is a pro-inflammatory chemokine receptor which, upon binding with its ligand CCL-2, promotes effector T-cell activation^10^. Non-obese diabetic (NOD) *CCR2*-/-mice develop T1D at a slower rate than NOD wild-type mice^11^. Serum CCL2 levels are different between T1D and controls in case-control studies; however, the direction of difference is contested^12-14^. Lower serum levels of CCL-2 have been reported in children with persistent islet autoantibodies compared to controls^15^, suggesting a potential causative role of *CCR2* in T1D pathogenesis^16,17^. *CCR2* has also been implicated in other immune diseases, including rheumatoid arthritis (RA)^18-20^ and multiple sclerosis (MS)^21-23^. However, clinical trials of CCR2 antagonists did not show reduce disease progression in either RA or MS patients^24,25^.

CCR2 inhibition is associated with increased serum levels of its ligand, CCL2 or MCP-1, in both mouse and human studies^24,26,27^. CCR2 activation by CCL2 has been shown to cellular uptake and internalization of the CCR2/CCL2 complex^28^. A recent GWAS identified SNPs near *CCR2* in the 3p21.31 genetic locus associated with serum CCL2 levels. These studies suggest that *cis*-acting genetic variants which alter CCR2 expressivity could paradoxically change serum CCL2 levels^29^.

In this report, the direction of the association between CCL2 level and T1D progression is determined in a longitudinal cohort (DAISY) and potential causal variants in the 3p21.31 locus were identified.

## RESULTS

### Serum CCL-2 levels decrease over time in DAISY participants who develop T1D compared to controls

In this study, we measured CCL-2 levels in longitudinally collected serum samples from 310 children at high risk for T1D participating in the Diabetes Autoimmunity Study in the Young (DAISY)^30,31^. At the time of analysis, 48 DAISY participants developed persistent islet autoantibodies and progressed to T1D (progressors) and an additional 42 participants developed persistent islet autoantibodies but have not progressed to T1D (non-progressors). Their longitudinal CCL-2 levels were compared to those of 220 islet autoantibody negative children (controls) similar to the cases in terms of age, sex, family history of T1D and HLA genotypes (**Table 1)**.

**Table 1.** Characteristics of study participants by study group. Categorical variables were analyzed using Pearson χ2 tests. Continuous variables were tested using the t test for differences in means or the Wilcoxon rank sum test for differences in medians

| **Table 1. Characteristics of study participants by study group.** |  |  |  |  |  |
| --- | --- | --- | --- | --- | --- |
| Categorical variables were analyzed using Pearson $\chi^2$ tests. Continuous variables were tested using the t test for differences in means or the Wilcoxon rank sum test for differences in medians. | | | | | |
|  |  | **Controls**<br>**N=220** | **Non-**<br>**progressors**<br>**N=42** | **Progressors**<br>**N=48** | **P value** |
| **Age of the first sample,**<br>**median (IQR)** |  | 1.3 (0.8-2.4) | 1.3 (0.8-3.1) | 1.3 (0.8-2.6) | 0.68 |
| **Number of serum samples,**<br>**median** |  | 7 (5-10) | 11 (7-16) | 14 (10-17) | <0.0001 |
| **HLA genotype, N**<br>**(%)** | **DR3/3 or**<br>**DR3/X or**<br>**DRX/X** | 89 (40.0) | 13 (31.0) | 12 (25.0) | 0.09 |
|  | **DR3/4 or**<br>**DR4/4 or**<br>**DR4/X** | 131 (60.0) | 29 (69.0) | 36 (75.0) |  |
| **Fist-degree**<br>**relative, N (%)** | **No** | 94 (42.7) | 16 (38.1) | 17 (35.4) | 0.60 |
|  | **Yes** | 126 (57.3) | 26 (61.9) | 31 (64.6) |  |
| **Sex, N (%)** | **Female** | 96 (43.6) | 22 (52.4) | 24 (50.0) | 0.48 |
|  | **Male** | 124 (56.4) | 20 (47.6) | 24 (50.0) |  |

The subjects were followed for 16.80 years on average (range 1.98 to 31.00 years). On average, 9.34 (range 3 to 30) serial serum samples were collected from each subject. The mean (SD) age at the appearance of islet autoantibodies was 4.59± 3.25 years in progressors and 7.54± 4.24 years in non-progressors. The mean age of T1D diagnosis in progressors was 12.22+ 5.68 years.

CCL-2 was detectable in all samples from the 310 DAISY subjects. CCL-2 levels decreased with age in all groups. We next tested whether the average CCL-2 levels differed among the three study groups, using linear mixed models adjusting for age, sex, HLA genotype, FDR status (**Table 2**). CCL-2 levels were significantly lower in both progressors (p=0.004) and non-progressors (p=0.005), compared to controls (**Supplemental Figure 1**). Subjects with high-risk HLA genotype (DR3/4 or DR4/4 or DR4/X) had lower MCP1 level compared to subjects with moderate-risk HLA genotype DR3/3 or DR3/X or DRX/X (p=0.009). However, CCL-2 levels did not differ by sex or by the presence of a first-degree relative.

**Table 2.** Multivariate mixed models analysis of the CCL-2 levels.

| **Table 2. Multivariate mixed models analysis of the CCL-2 levels** |  |  |  |  |
| --- | --- | --- | --- | --- |
| Effect |  | Estimate | Standard Error | Pr > \|t\| |
| Intercept |  | 89.63 | 2.26 | <.0001 |
| Age*Age |  | -0.03 | 0.01 | **0.0013** |
| Sex | Female vs Male | -0.34 | 1.85 | 0.85 |
| HLA genotype | DR3/4 or DR4/4 or DR4/X vs DR3/3 or DR3/X or DRX/X | -5.20 | 2.00 | **0.009** |
| First-degree relative with T1D | Yes vs No | 0.68 | 1.95 | 0.73 |
| Study Group | Non-progressors vs Controls | -7.32 | 2.62 | **0.005** |
|  | Progressors vs Controls | -7.07 | 2.46 | **0.004** |

**Figure 1.**
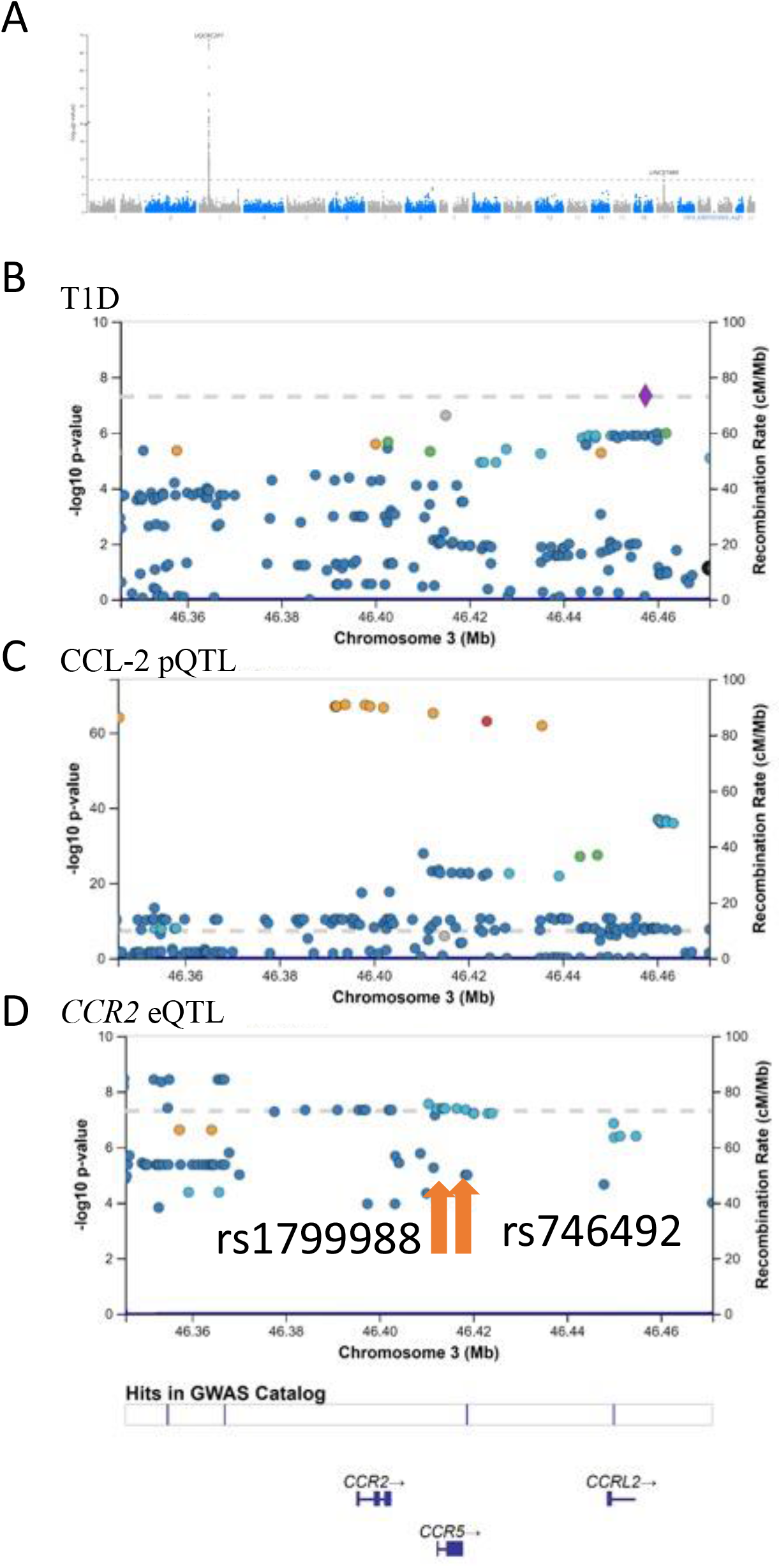
GWAS for CCL-2 serum levels and fine mapping for type 1 diabetes. A) Manhattan plot of the serum CCL-2 protein quantitative trait loci (pQTL) from Folkersen et al^29^. Data were downloaded from Xenodo and plotted using the LocusZoom plot package. The genome wide significant SNPs are from the 3p21.31 region. B) LocusZoom plot of SNPs significance on T1D association testing from T1DGC fine mapping study. C) LocusZoom plot of SNPs significance on CCL-2 pQTL from Figure 1A. D) LocusZoom plot of SNPs significance on CCR2 eQTL from GTEx. The potential causal variants rs1799988 (chr3:46412259) and rs746492 (chr3:46417312) are indicated with arrows.

### The 3p21.31 genetic locus is associated with serum CCL-2 levels

Genetic variants associated with serum CCL-2 levels were determined by downloading summary statistics from a study estimating cis- and trans-protein quantitative trait loci (pQTLs) for over 600 proteins using over 20,000 subjects ^29^. One locus contributing to serum CCL-2 levels at 3p21.31 was identified **(Figure 1A)**, centered around the *CCR2* gene **(Figure 1B)**. The SNPs on chromosome 3 accounted for ∼10% of the variation in CCL-2 serum level, which explains 70% of estimated heritability^29^. An additional study in the GWAS catalog^32^ validated the association between the 3p21.31 locus and serum CCL-2 levels^33^.

### Potential causal variants for serum CCL-2 level regulation and T1D pathogenesis

Since the 3p21.31 locus is associated with both CCL-2 serum levels and T1D, and CCL-2 serum levels are associated with T1D, genetic variants associated with T1D through regulatory control of CCL-2 serum levels were targeted for evaluation. Multiple levels of data were integrated to identify a potential set of causal variants from the 3p21.31 locus. Summary statistics were obtained from the T1DGC ImmunoChip fine mapping data^6^, which profiled 135,870 custom selected SNPs on 18,892 subjects. After merging the summary statistics from CCL-2 protein quantitative trait loci (pQTL) and T1DGC data, a total of 8,804 SNPs on chromosome 3 overlapped both the CCL-2 and T1DGC studies, including 576 SNPs between position chr3:46,300,000 - 46,800,000 (**Figure 1C**). Using a Bonferroni-adjusted p-value cut-off, 225 SNPs were associated (p < 8.7 x10^−5^) with CCL-2 serum levels, with 63 SNPs associated with T1D. A total of 47 SNPs were associated with both T1D and CCL-2 serum levels. Expression QTLs (eQTLs) associated with *CCR2* were obtained from the Genotype-Tissue Expression (GTEx) portal for whole blood. Eighty-six SNPs overlapped the combined CCL-2 pQTL and T1DGC fine mapping data and the GTEx whole blood eQTL data at chr 3:46,300,000 - 46,800,000 (**Figure 1D**).

Two SNPs, rs1799988 and rs746492, were significantly associated with T1D, CCL2 serum pQTL, and *CCR2* eQTLs (**Figure 2**), suggesting that the two SNPs could play a regulatory role in CCR2/CCL2 pathway activation and progression to T1D. We therefore propose a mechanism linking the two potential causal variants to T1D pathogenesis. Individuals with these variants have associated increased immune cell expression of *CCR2* and the associated increase in CCR2/CCL2 signaling, which causes immune-mediated islet destruction and depletion of serum CCL-2 levels **(Figure 3)**.

**Figure 2.**
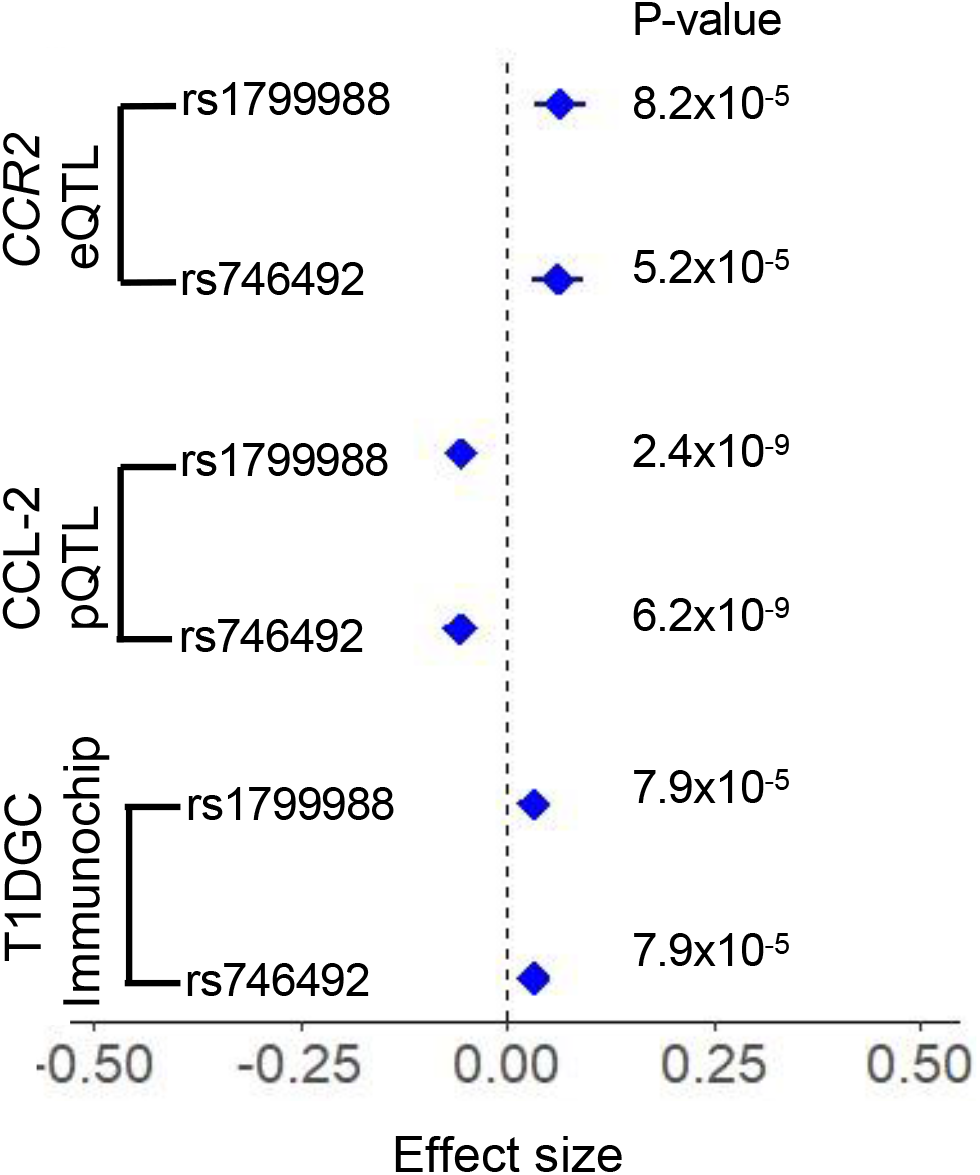
Forest plot of potential causal SNPs (rs1799988 and rs746492) *CCR2* eQTL, CCL2 pQTL, and T1D Immunochip Association. The axis represents effect size with bars showing 95% confidence interval.

**Figure 3.**
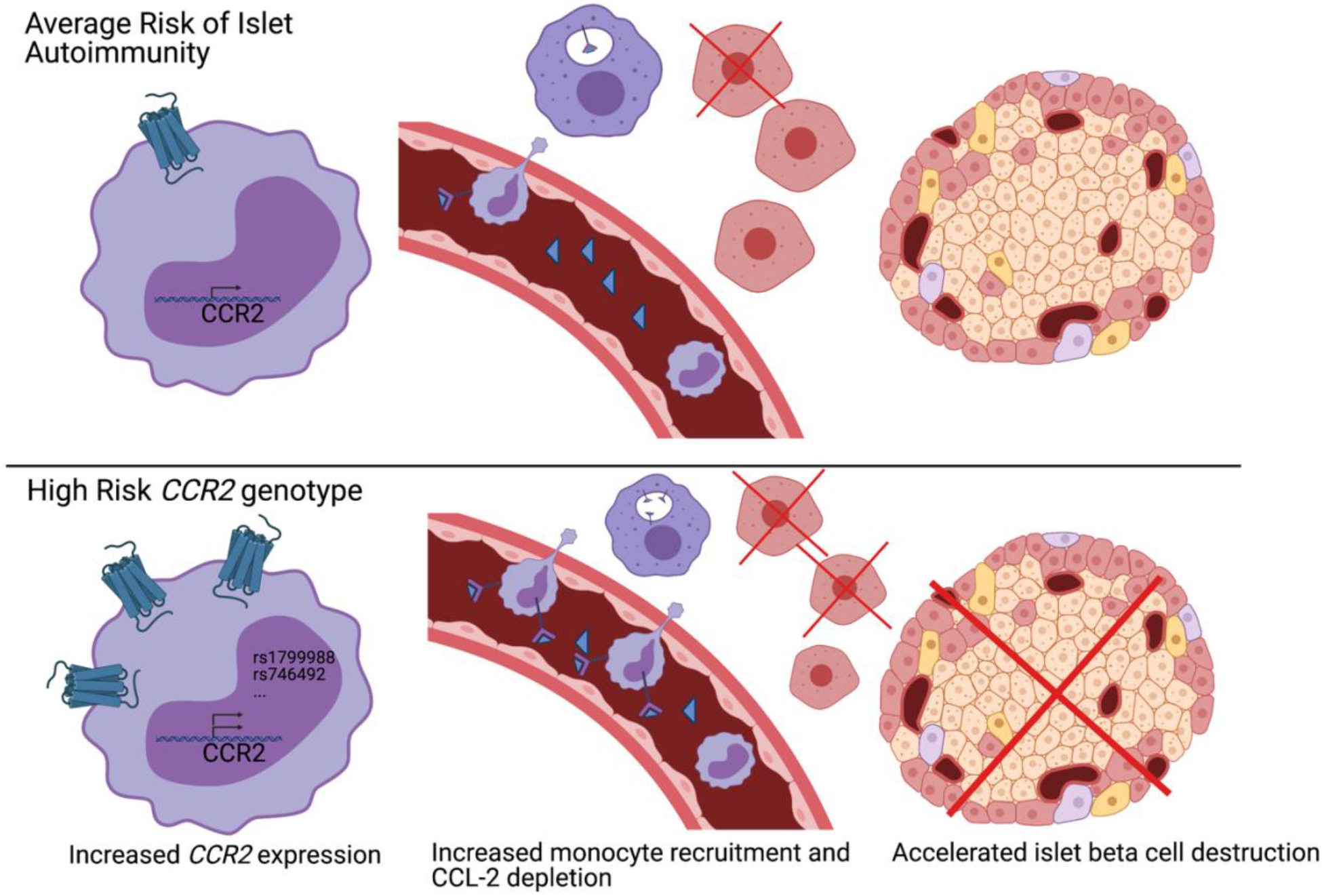
Proposed model of 3p21.31 locus influence on progression to type 1 diabetes. Individuals with high risk *CCR2* genotypes are more prone to increased *CCR2* expression, increased CCR2/CCL2 axis signaling with subsequent depletion of serum CCL2, increased monocyte recruitment, and increased rate of pancreatic islet destruction, resulting in an increased rate of progression to type 1 diabetes. Created with BioRender.com.

## DISCUSSION

While some studies have shown that serum CCL-2 levels are increased in T1D subjects compared to normal controls, our previous work and current findings do not agree with this^12-14^. By analyzing serum CCL-2 levels in a longitudinal cohort, we were able to demonstrate the decrease in serum CCL-2 levels as subjects progressed from autoantibody positivity to T1D **(Table 2)**. The decrease in serum CCL-2 with disease progression is in agreement with our proposed mechanism. *Cis*-acting genetic variants increase *CCR2* expression above the average population level which increases CCR2/CCL2 axis signaling and monocyte recruitment to the pancreatic islet. The increased signaling depletes the serum pool of CCL2 **(Figure 3)**. These findings contradict the simple logic that increased serum inflammatory markers indicate a pro-inflammatory state. A similar mechanism has been described where decreased IL-6 serum levels are associated with worse cardiovascular prognosis due to increased IL-6R/IL-6 signaling in the vascular wall^34,35^. Our findings provide an additional example of decreased serum levels of a pro-inflammatory molecule associated with an increased inflammatory state.

Interestingly, serum CCL2 levels were more likely to be elevated at 3-4 months in infants who will develop islet autoimmunity in The Environmental Determinants of Diabetes in the Young^36^ (TEDDY) high risk genetic cohort (M. Rewers, personal communication). This may be due to early enterovirus infection, which has been shown to increase type 1 interferon and CCL2 expression^37^. A lack of statistical significance of decreased CCL2 levels in progression to islet autoimmunity and type 1 diabetes may be due to lack of statistical power in this cohort since 1) less samples were collected after seroconversion and 2) the cohort is a genetically pre-select group which may lack the traditional variation in CCL2 levels.

We were able to narrow the list of potential causal variants by integrating the CCL-2 pQTL and T1D Immunochip fine mapping. Although this integration provides high confidence for these SNPs, they will require functional validation. Additionally, while the T1D fine mapping study used a custom SNP array for autoimmune diseases^38^, the CCL-2 GWAS and the GTeX eQTL study^39^ used a broader SNP array. Thus, SNPs which were not assessed in all SNP arrays could not be fairly evaluated and may still regulate the CCR2/CCL2 axis.

Our findings support the potential of CCR2 inhibition in delaying the progression of islet autoantibody positive individuals to T1D. Several CCR2 inhibitors have been tested in clinical trials for cardiovascular disease, diabetic nephropathy, cancer, and autoimmunity^40-44^. Within autoimmunity, CCR2 inhibition did not change disease progression for subjects with rheumatoid arthritis or multiple sclerosis. One potential explanation is that CCR2 inhibition was targeted too late in the disease course and that earlier targeting based on serology and before clinical manifestations would modulate clinical progression^45^. Additionally, individuals with high CCL-2 serum levels, indicating a normal or low expression and activity of CCR2 would likely not benefit as much as individuals with high *CCR2* expression and activity with lower CCL-2 serum levels. Thus, potential future trials for T1D prevention should target autoantibody positive individuals with lower serum levels of CCL-2.

In conclusion, we mapped the T1D association of the 3p21.31 locus to the *CCR2* gene. Increased *CCR2* gene expression decreases serum levels of CCL-2 and is associated with increased immune destruction of pancreatic islets and quicker progression to T1D.

## METHODS

### Human subjects

The prospective cohort was part of the DAISY study^30,31^. The sex, age, family history of T1D, and HLA-DQB1 genotypes for all subjects were summarized in **Table 1**. Serum samples were collected in clot activator tubes and processed and stored within less than 1 hour. Repeated freezer/thawing cycles were avoided for all samples. Islet autoantibodies to insulin, GAD, IA-2 and ZnT8 were measured by radio-binding (RBA) assay. Persistence was defined by presence of at least one of the autoantibodies on at least two consecutive visits 3-6 months apart.

### Laboratory assays

Serum levels of CCL-2 were measured using multiplex fluorescent immunoassays (Millipore, Billerica MA) according to the manufacturer’s recommendations. The coefficients of variation (CV, <10%) and the minimum detectable dose (MDD, CCL-2: 0.9) for the assays were provided by the manufacturer with the assay kits.

Serum samples (5x diluted) were incubated with capture antibodies immobilized on polystyrene beads for one hour. The beads were then washed and further incubated with biotinylated detection antibody cocktail for one hour. Beads were washed twice to remove unbound detection antibody and then incubated with phycoerythrin-labeled streptavidin for thirty minutes. Prior to reading, the beads were washed 2 times and suspended in 60µl of wash buffer. The median fluorescence intensities (MFI) were measured on a FlexMAP 3D array reader (Millipore, Billerica, MA) using the following instrument settings: events/bead: 50, minimum events: 0, flow rate: 60µl/min, Sample size: 50ul and discriminator gate: 8000-13500. Before profiling, the serum dilutions were optimized by performing the assays at different serum dilutions to ensure that the majority of the data falls within the linear range of the standard curve.

Protein concentrations were estimated using a regression fit to the standard curve with known concentration included on each plate using a serial dilution series. The data were subjected to several quality control steps before further analysis^12^.

### Data acquisition

DAISY phenotype data was provided by MR. CCL-2 pQTL data were downloaded from Zenodo (https://zenodo.org/record/2615265#.YGs9GEhKjYk) on 05/01/2021. T1DGC summary statistics data were provided by SOG and SR^6^. CCR2 eQTL data were downloaded from GTEx portal (https://www.gtexportal.org/home/gene/CCR2) on 05/01/2021.

### Statistical analysis

Statistical analyses of serum CCL2 levels were performed using SAS software version 9.4. A p-value less than 0.05 was considered significant. Because CCL-2 values are not normally distributed, Box-Cox transformation was performed to normalize the data. The optimal power parameter (lamda) was obtained using PROC TRANSREG by using a maximum likelihood criterion^46^. The optimal power parameter (Lamda) was found to be 0.75 and tCCL-2=((CCL-2)**0.75-1)/0.75. Linear mixed models (PROC mixed) are used to compare the difference in Box-Cox transformed CCL-2 levels among the three study groups, adjusting for age, gender (female vs male) and HLA genotype ([DR3/4 or DR4/4 or DR4/X] vs [DR3/3 or DR3/X or DRX/X]) and FDR status (FDR vs GP) with random intercept in the model^47^. Fit statistics between different curves over age were compared for a lower AIC/BIC and the model with quadratic curve over age was chosen as the final model.

Additional statistical analyses were performed using the R language and environment for statistical computing (R version 4.0.5; R Foundation for Statistical Computing; www.r-project.org). PLINK1.9^48^ was used for pre-processing the genotype data. LocusZoom^49^ was used to generate both Manhattan and LocusZoom plots. All p-values were two–tailed and a p< 0.05 after adjusting for multiple testing was considered statistically significant.

## Data Availability

The data that support the findings from this study are openly available from Zenodo (https://zenodo.org/record/2615265#.YGs9GEhKjYk) and the GTEx portal (https://www.gtexportal.org/home/gene/CCR2). The T1DGC Immunochip summary statistics data and the CCL-2 serum DAISY data are available by request to the authors.

## Acknowledgements

Support for this work to JXS was provided by the National Institutes of Health ([R21HD050196](https://public.era.nih.gov/grantfolder/piAppDetails/genericStatus.do?encryptedParam=(v2)ETMsDgAAAWDgE1r-ABRBRVMvQ0JDL1BLQ1M1UGFkZGluZwCAABAAEMwuYtIFSDk0YfK-Nuphb90AAAAgAcuKccRAZX9Zf6uYMr6TpbwEibMk2IFnjXgZPZ6CbIsAFP1BxIWLl3WiLLbVVdQJJof8dBPG), R33HD050196, and 2RO1HD37800). SP was supported by Postdoctoral Fellowship (10-2006-792 and 3-2004-195) and Career Development Award (2-2011-153) from JDRF. PMHT was supported by NIH/NIDDK fellowship (F30DK121461). Diabetes Autoimmunity Study in the Young (MR, KW, and FD) is supported by the National Institutes of Health (R01 DK032493 and P30 DK116073) and Helmsley Charitable Trust (G-1901-03687). This research utilizes resources provided by the Type 1 Diabetes Genetics Consortium, a collaborative clinical study sponsored by the National Institute of Diabetes and Digestive and Kidney Diseases (NIDDK), National Institute of Allergy and Infectious Diseases (NIAID), National Human Genome Research Institute (NHGRI), National Institute of Child Health and Human Development (NICHD), and JDRF and supported by U01 DK062418.

All authors declare no conflict of interest.

## Author contributions

PMHT wrote the manuscript and researched data. SP acquired the Luminex data; SP, KBS, EK, PMHT, FD, KW and MR were responsible for data analysis. DH, KW, MR and JXS contributed to clinical samples. S O-G and SSR generated fine mapping data. All authors contributed to writing and editing of the manuscript.

---

**Supplemental Fig. 1.**
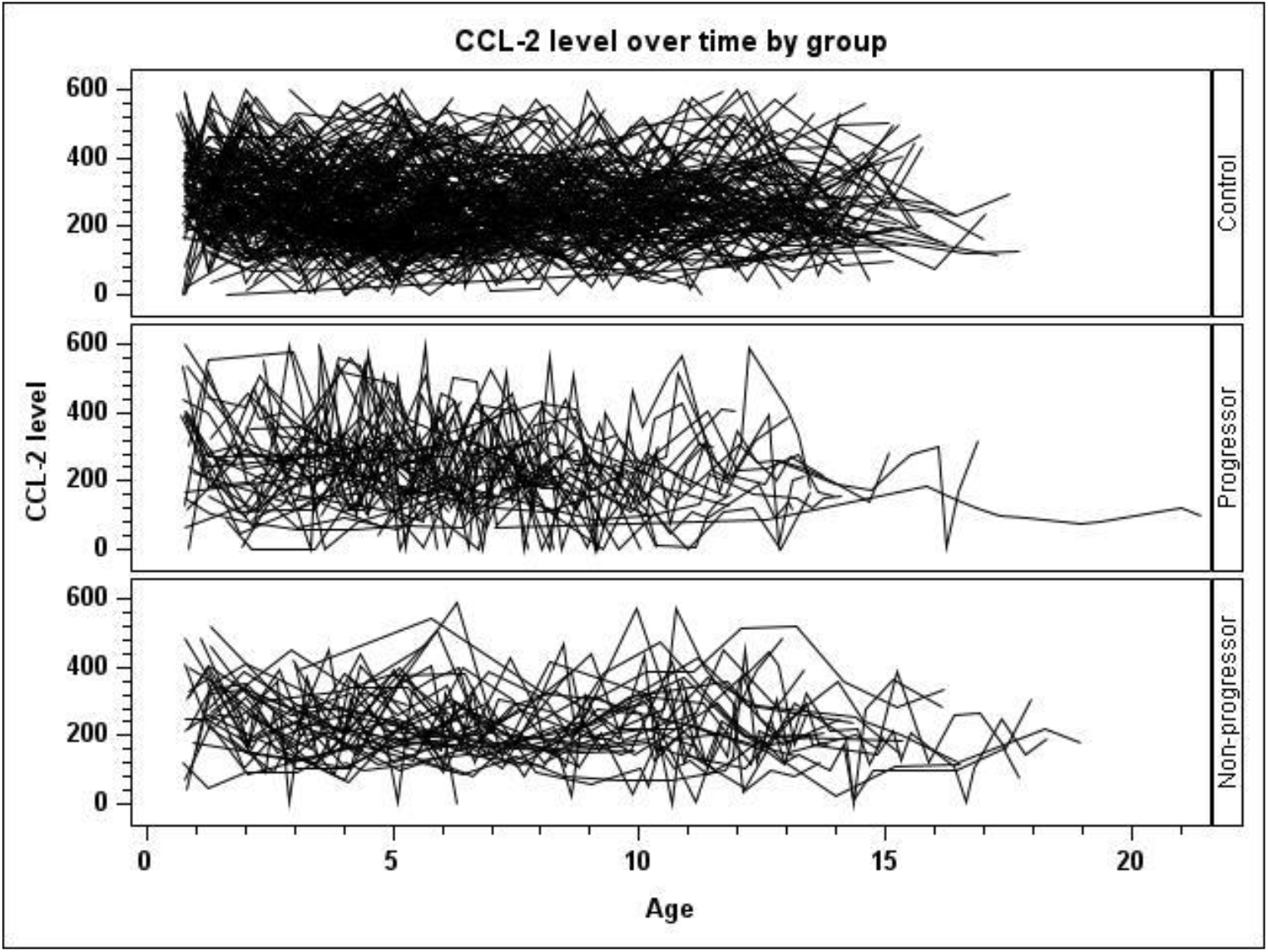
The following graph is spaghetti plot of the CCL-2 level over age among different groups with total number of subjects=310.

**Supplemental Fig 2.**
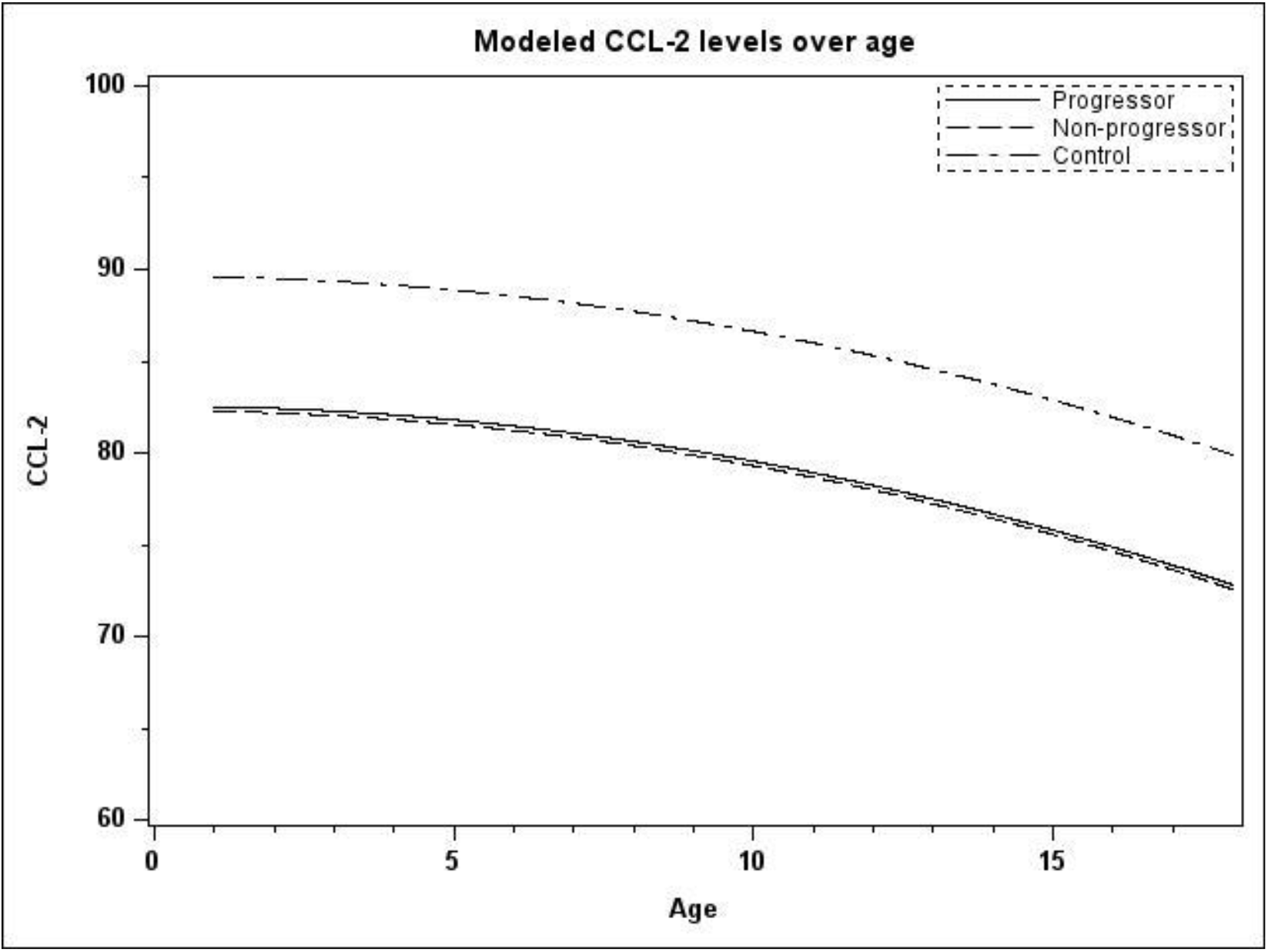
Modeled CCL-2 levels for controls (upper curve), progressors, and non-progressor from the mixed model with a quadratic curve over age. Fit computed for male and HLA group of DR3/3 or DR3/X or DRX/X and GP.

